# Higher dose corticosteroids in hospitalised COVID-19 patients with hypoxia but not requiring ventilatory support (RECOVERY): a randomised, controlled, open-label, platform trial

**DOI:** 10.1101/2022.12.16.22283578

**Authors:** RECOVERY Collaborative Group, Peter W Horby, Jonathan R Emberson, Buddha Basnyat, Mark Campbell, Leon Peto, Guilherme Pessoa-Amorim, Natalie Staplin, Raph L Hamers, John Amuasi, Jeremy Nel, Evelyne Kestelyn, Manisha Rawal, Roshan Kumar Jha, Nguyen Thanh Phong, Uun Samardi, Damodar Paudel, Pham Ngoc Thach, Nasronudin Nasronudin, Emma Stratton, Louise Mew, Rahul Sarkar, J Kenneth Baillie, Maya H Buch, Jeremy Day, Saul N Faust, Thomas Jaki, Katie Jeffery, Edmund Juszczak, Marian Knight, Wei Shen Lim, Marion Mafham, Alan Montgomery, Andrew Mumford, Kathryn Rowan, Guy Thwaites, Richard Haynes, Martin J Landray

## Abstract

**Background:** Low-dose corticosteroids have been shown to reduce mortality for hypoxic COVID-19 patients requiring oxygen or ventilatory support (non-invasive mechanical ventilation, invasive mechanical ventilation or extra-corporeal membrane oxygenation). We evaluated the use of a higher dose of corticosteroids in this patient group.

**Methods:** This randomised, controlled, open-label platform trial (Randomised Evaluation of COVID-19 Therapy [RECOVERY]) is assessing multiple possible treatments in patients hospitalised for COVID-19. Eligible and consenting adult patients with clinical evidence of hypoxia (i.e. receiving oxygen or with oxygen saturation <92% on room air) were randomly allocated (1:1) to either usual care with higher dose corticosteroids (dexamethasone 20 mg once daily for 5 days followed by 10 mg once daily for 5 days or until discharge if sooner) or usual standard of care alone (which includes dexamethasone 6 mg once daily for 10 days or until discharge if sooner). The primary outcome was 28-day mortality. On 11 May 2022, the independent Data Monitoring Committee recommended stopping recruitment of patients receiving no oxygen or simple oxygen only to this comparison due to safety concerns. We report the results for these participants only. Recruitment of patients receiving ventilatory support continues. The RECOVERY trial is registered with ISRCTN (50189673) and clinicaltrials.gov (NCT04381936).

**Findings:** Between 25 May 2021 and 12 May 2022, 1272 COVID-19 patients with hypoxia and receiving no oxygen (1%) or simple oxygen only (99%) were randomly allocated to receive usual care plus higher dose corticosteroids versus usual care alone (of whom 87% received low dose corticosteroids during the follow-up period). Of those randomised, 745 (59%) were in Asia, 512 (40%) in the UK and 15 (1%) in Africa. 248 (19%) had diabetes mellitus. Overall, 121 (18%) of 659 patients allocated to higher dose corticosteroids versus 75 (12%) of 613 patients allocated to usual care died within 28 days (rate ratio [RR] 1·56; 95% CI 1·18-2·06; p=0·0020). There was also an excess of pneumonia reported to be due to non-COVID infection (10% vs. 6%; absolute difference 3.7%; 95% CI 0.7-6.6) and an increase in hyperglycaemia requiring increased insulin dose (22% vs. 14%; absolute difference 7.4%; 95% CI 3.2-11.5).

**Interpretation:** In patients hospitalised for COVID-19 with clinical hypoxia but requiring either no oxygen or simple oxygen only, higher dose corticosteroids significantly increased the risk of death compared to usual care, which included low dose corticosteroids. The RECOVERY trial continues to assess the effects of higher dose corticosteroids in patients hospitalised with COVID-19 who require non-invasive ventilation, invasive mechanical ventilation or extra-corporeal membrane oxygenation.

**Funding:** UK Research and Innovation (Medical Research Council) and National Institute of Health and Care Research (Grant ref: MC_PC_19056), and Wellcome Trust (Grant Ref: 222406/Z/20/Z).

## INTRODUCTION

The RECOVERY trial has previously shown that the use of corticosteroids (using dexamethasone 6 mg once daily for ten days or until discharge if sooner) reduces the risk of death in patients admitted to hospital with COVID-19 and hypoxia.^1^ Subsequent findings that additional immunosuppression with an interleukin-6 (IL-6) receptor blocker and/or a Janus kinase (JAK) inhibitor further reduces the risk of death in these patients raises the question whether simply increasing the dose of corticosteroid rather than adding other immunomodulators could confer additional benefits at substantially lower cost.^2,3^

Initially, the dose of dexamethasone chosen for the RECOVERY trial was 6 milligrams once daily on the basis that there is considerable experience of using similar doses of corticosteroids safely for exacerbations of asthma and COPD.^4,5^ However, the optimal corticosteroid dose in COVID-19 treatment has not been determined. Higher doses of corticosteroids have been used in non-COVID acute respiratory distress syndrome (ARDS) and in other COVID-19 clinical trials with reported clinical benefit, but these trials have been restricted to severe or critically ill patients and have not directly compared higher doses with the lower dose used in RECOVERY.^6-11^ A WHO meta-analysis of randomised controlled trials in critically ill COVID-19 indicated similar mortality benefit with lower and higher-dose corticosteroids, but estimates were imprecise.^9^

In April 2021, the United Kingdom COVID-19 Therapeutics Advisory Panel recommended that the RECOVERY trial study higher dose of corticosteroids.^12^ The RECOVERY trial therefore established a randomised evaluation of the effects of higher-dose corticosteroids versus usual care for adult patients who had been admitted to hospital with COVID-19 and had clinical evidence of hypoxia. Usual care for hypoxic COVID-19 patients includes low dose corticosteroids. On 11 May 2022, the independent Data Monitoring Committee recommended that this comparison be halted for those patients receiving no oxygen or simple oxygen only on the grounds of safety. Here we report the results for these patients. Recruitment to this comparison continues as planned for patients receiving non-invasive ventilation or invasive mechanical ventilation.

## METHODS

### Study design and participants

The Randomised Evaluation of COVID-19 therapy (RECOVERY) trial is an investigator-initiated, individually randomised, controlled, open-label, platform trial to evaluate the effects of potential treatments in patients hospitalised with COVID-19. Details of the trial design and results for other possible treatments (dexamethasone, hydroxychloroquine, lopinavir-ritonavir, azithromycin, tocilizumab, convalescent plasma, colchicine, aspirin, casirivimab plus imdevimab, and baricitinib) have been published previously.^1-3,13-19^ The trial is underway at hospital organisations in the United Kingdom, supported by the National Institute for Health Research Clinical Research Network, as well as in South and Southeast Asia and Africa. Of these, 81 hospitals in the UK, 5 in Nepal, 2 in Indonesia, 2 in Vietnam, 2 in South Africa, and 1 in Ghana enrolled participants in the evaluation of higher dose corticosteroids (appendix pp 2-31). The trial is coordinated by the Nuffield Department of Population Health at the University of Oxford (Oxford, UK), the trial sponsor. The trial is conducted in accordance with the principles of the International Conference on Harmonisation–Good Clinical Practice guidelines. The protocol is approved by all relevant regulatory authorities and ethics committees in each participating country (appendix pp 32) The protocol and statistical analysis plan are available in the appendix (pp 71-152) with additional information available on the study website www.recoverytrial.net.

Patients aged at least 18 years admitted to hospital were eligible for the study if they had clinically suspected or laboratory confirmed SARS-CoV-2 infection, clinical evidence of hypoxia (i.e. receiving oxygen with or without other forms of respiratory support, or with oxygen saturations <92% on room air) and no medical history that might, in the opinion of the attending clinician, put the patient at significant risk if they were to participate in the trial. Patients were ineligible for the comparison of higher dose corticosteroid vs. usual care if there was a known contra-indication to short-term use of corticosteroids, suspected or confirmed influenza, or current use of nirmatrelvir/ritonavir, ritonavir or other potent CYP3A inhibitors. Endemic infections were screened for as required by local practice. Other immunomodulatory therapies were not contra-indicated but investigators were advised to consider the total burden of such therapy (e.g. combining an IL-6 receptor antagonist with higher dose corticosteroid). Written informed consent was obtained from all patients, or a legal representative if patients were too unwell or unable to provide consent.

### Randomisation and masking

Baseline data were collected using a web-based case report form that included demographics, level of respiratory support, major comorbidities, suitability of the study treatment for a particular patient, SARS-CoV-2 vaccination status, and treatment availability at the study site (appendix pp 44-47).

Eligible and consenting patients were assigned in a 1:1 ratio to either usual standard of care plus higher dose corticosteroids or usual standard of care alone (which includes low dose corticosteroids, usually dexamethasone 6 mg once daily for 10 days or until discharge if sooner), using web-based simple (unstratified) randomisation with allocation concealed until after randomisation (appendix pp 42-44).^20,21^ Patients allocated to higher dose corticosteroid were to receive dexamethasone 20 mg daily for 5 days followed by dexamethasone 10 mg for 5 days (or until discharge if sooner). Alternative corticosteroid regimens for pregnant women are described in the protocol (appendix p 78).

As a platform trial, and in a factorial design, patients could be simultaneously randomised to other treatment groups: i) empagliflozin versus usual care, ii) sotrovimab versus usual care, and iii) molnupiravir versus usual care. Further details of when these factorial randomisations were open are provided in the supplementary appendix (p 42-44). Participants and local study staff were not masked to the allocated treatment. The Trial Steering Committee, investigators, and all other individuals involved in the trial are masked to outcome data while recruitment and 28-day follow-up are ongoing.

### Procedures

An online follow-up form was completed by site staff when patients were discharged, had died, or at 28 days after randomisation, whichever occurred first (appendix pp 48-56). Information was recorded on adherence to allocated trial treatment, receipt of other COVID-19 treatments, duration of admission, receipt of respiratory or renal support, new cardiac arrhythmia, thrombosis, clinically significant bleeding, non-COVID infection, metabolic complications (collected from 28 July 2021 onwards), and vital status (including cause of death). In addition, in the UK, routinely collected healthcare and registry data were obtained, including information on vital status at day 28 (with date and cause of death); discharge from hospital; and receipt of respiratory support or renal replacement therapy. For sites outside the UK a further case report form (appendix p 58) collected vital status at day 28 (if not already reported on follow-up form).

### Outcomes

Outcomes were assessed at 28 days after randomisation, with further analyses specified at 6 months. The primary outcome was 28-day all-cause mortality. Secondary outcomes were time to discharge from hospital, and, among patients not on invasive mechanical ventilation at randomisation, the composite outcome of invasive mechanical ventilation (including extra-corporeal membrane oxygenation) or death. Prespecified subsidiary clinical outcomes were use of invasive or non-invasive ventilation among patients not on any ventilation at randomisation, and use of renal dialysis or haemofiltration among patients not receiving such treatment at randomisation. Prespecified safety outcomes were cause-specific mortality, major cardiac arrhythmia, thrombotic and major bleeding events, other infections and metabolic complications. Information on suspected serious adverse reactions was collected in an expedited fashion to comply with regulatory requirements. Details of the methods used to ascertain and derive outcomes are provided in the appendix (pp.153-176).

### Sample size and role of the independent Data Monitoring Committee

As stated in the protocol, appropriate sample sizes could not be estimated when the trial was being planned. However, the intention for this comparison was to continue recruitment until sufficient primary outcomes had accrued to have 90% power to detect a proportional risk reduction of 20% at 2P=0.01.

The independent Data Monitoring Committee reviewed unblinded analyses of the study data and any other information considered relevant to the trial at intervals of around 2-3 months (depending on speed of enrolment) and was charged with determining if, in their view, the randomised comparisons in the study provided evidence on mortality that was strong enough (with a range of uncertainty around the results that was narrow enough) to affect national and global treatment strategies (appendix p 59).

On 11 May 2022, the Data Monitoring Committee recommended stopping recruitment to the higher dose corticosteroid comparison for patients who require no oxygen or simple oxygen only at randomisation due to safety concerns (appendix p 60). The Data Monitoring Committee encouraged continuing recruitment of all those patients who, at randomisation, require either non-invasive ventilation, invasive mechanical ventilation or ECMO.

On 13 May 2022, recruitment of patients on oxygen or simple oxygen only to this comparison was stopped, investigators were notified and advised not to administer further higher dose corticosteroids to this group as part of the trial. Follow-up of all patients continued as planned. The regulatory authorities and ethics committees in each participating country were notified. Trial procedures were changed as an urgent safety measure and the protocol was subsequently modified and approved accordingly. Recruitment and follow-up of patients receiving non-invasive ventilation or invasive mechanical ventilation remains open.

### Statistical Analysis

All analyses were limited to the subgroup of patients on no or simple oxygen at randomization. For all outcomes, intention-to-treat analyses compared patients randomised to higher dose corticosteroids with patients randomised to usual care. For the primary outcome of 28-day mortality, the log-rank observed minus expected statistic and its variance were used to both test the null hypothesis of equal survival curves (i.e., the log-rank test) and to calculate the one-step estimate of the average mortality rate ratio. We constructed Kaplan-Meier survival curves to display cumulative mortality over the 28-day period. We used the same method to analyse time to hospital discharge with patients who died in hospital right-censored on day 29. Median time to discharge was derived from Kaplan-Meier estimates. For the composite secondary outcome of progression to invasive mechanical ventilation or death within 28 days, and the subsidiary clinical outcomes of receipt of ventilation and use of haemodialysis or haemofiltration, the precise dates were not available and so the risk ratio was estimated instead. Estimates of rate and risk ratios (both denoted RR) are shown with 95% confidence intervals. For the primary outcome of 28-day mortality, a sensitivity analysis adjusted for age was done using Cox regression.

Since the analyses presented here relate only to the subset of participants who were on no oxygen or simple oxygen only at randomisation, any analyses of the primary outcome in further subgroups defined by different baseline characteristics must be considered exploratory in nature. With that caveat, we present analyses of the primary outcome by age, sex, ethnicity, days since symptom onset, and place of recruitment (UK, elsewhere) with tests of heterogeneity or trend, as appropriate. We have not presented analyses by subgroups for the secondary or other outcomes. Results for the pre-specified other clinical outcomes and safety outcomes are presented. The full database is held by the study team which collected the data from study sites and performed the analyses at the Nuffield Department of Population Health, University of Oxford (Oxford, UK). Analyses were performed using SAS version 9.4 and R version 4.0.3. The trial is registered with ISRCTN (50189673) and clinicaltrials.gov (NCT04381936).

### Role of the funding source

The funders of the study had no role in study design, data collection, data analysis, data interpretation, or writing of the report. The corresponding authors had full access to all the data in the study and had final responsibility for the decision to submit for publication.

## RESULTS

Recruitment to the evaluation of higher dose corticosteroids commenced on 25 May 2021 outside the UK and 29 December 2021 in the UK (following closure of the baricitinib comparison) and ended worldwide for those on no oxygen or simple oxygen on 13 May 2022 following the Data Monitoring Committee recommendation.^3^ Of 1680 patients enrolled in this comparison during this period, 1272 patients on no oxygen or simple oxygen only are included in this evaluation. Of these, 659 were randomly allocated to higher dose corticosteroids and 613 patients were randomly allocated to usual care (Figure 1). The mean age of these participants was 61.1 years (SD 17.5) (webtable 1) with those randomly allocated to higher dose corticosteroids on average 1.9 years younger than those allocated usual care group (Table 1). 745 (59%) were in Asia, 512 (40%) in the UK, and 15 (1%) in Africa. 248 (19%) had a history of diabetes mellitus, and 1264 (99%) were receiving simple oxygen (webtable 1).

**Figure 1:**
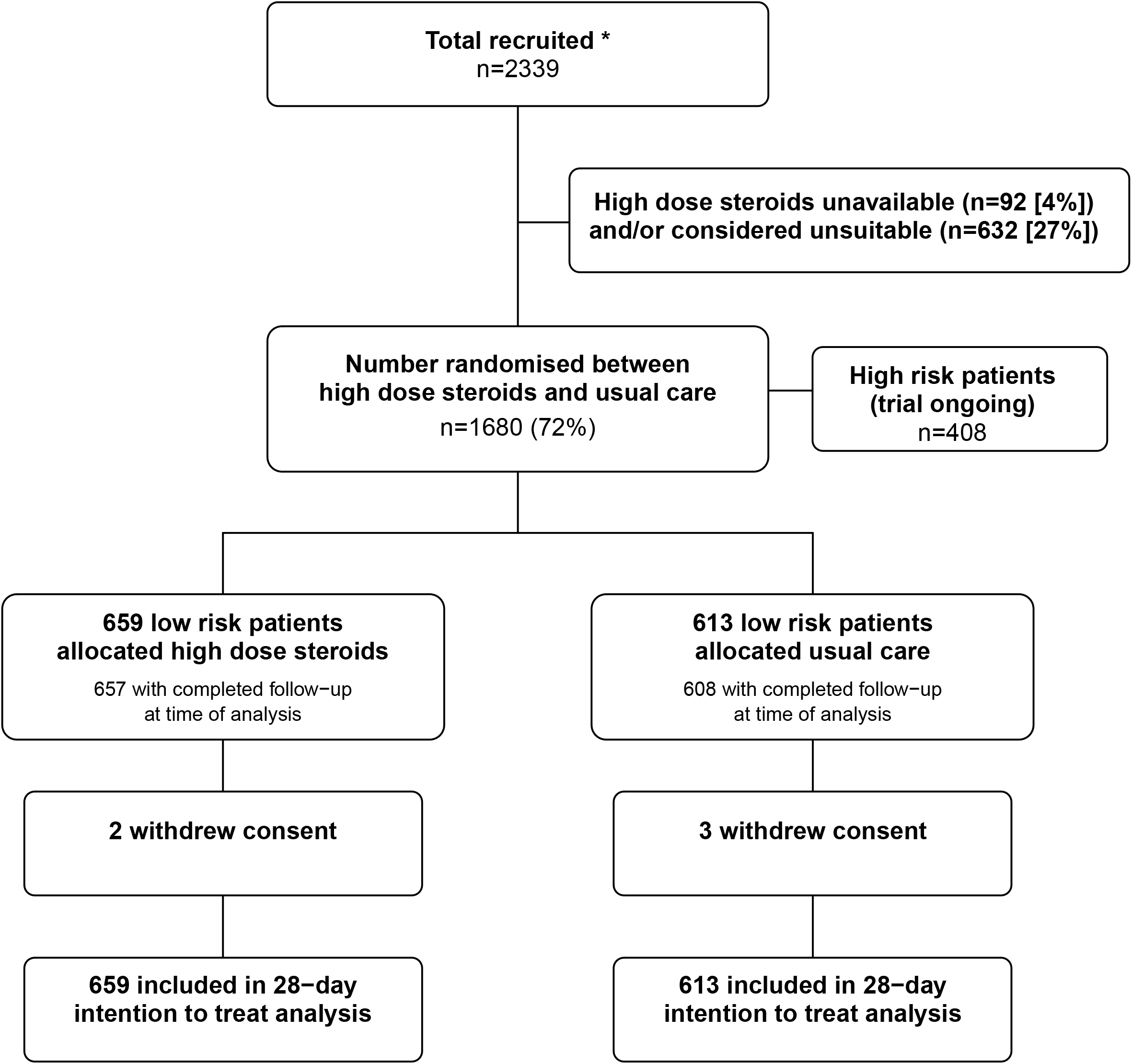
Trial profile. ITT=intention to treat. Higher dose corticosteroid unavailable and higher dose corticosteroid considered unsuitable are not mutually exclusive. High risk patients are those that were receiving non-invasive ventilation, invasive mechanical ventilation or ECMO at randomisation. Low risk patients are those that were hypoxic but receiving no oxygen or simple oxygen only. * Number recruited overall during period that participants on no oxygen or simple oxygen only could be recruited into the higher dose corticosteroid comparison.

**Table 1:**
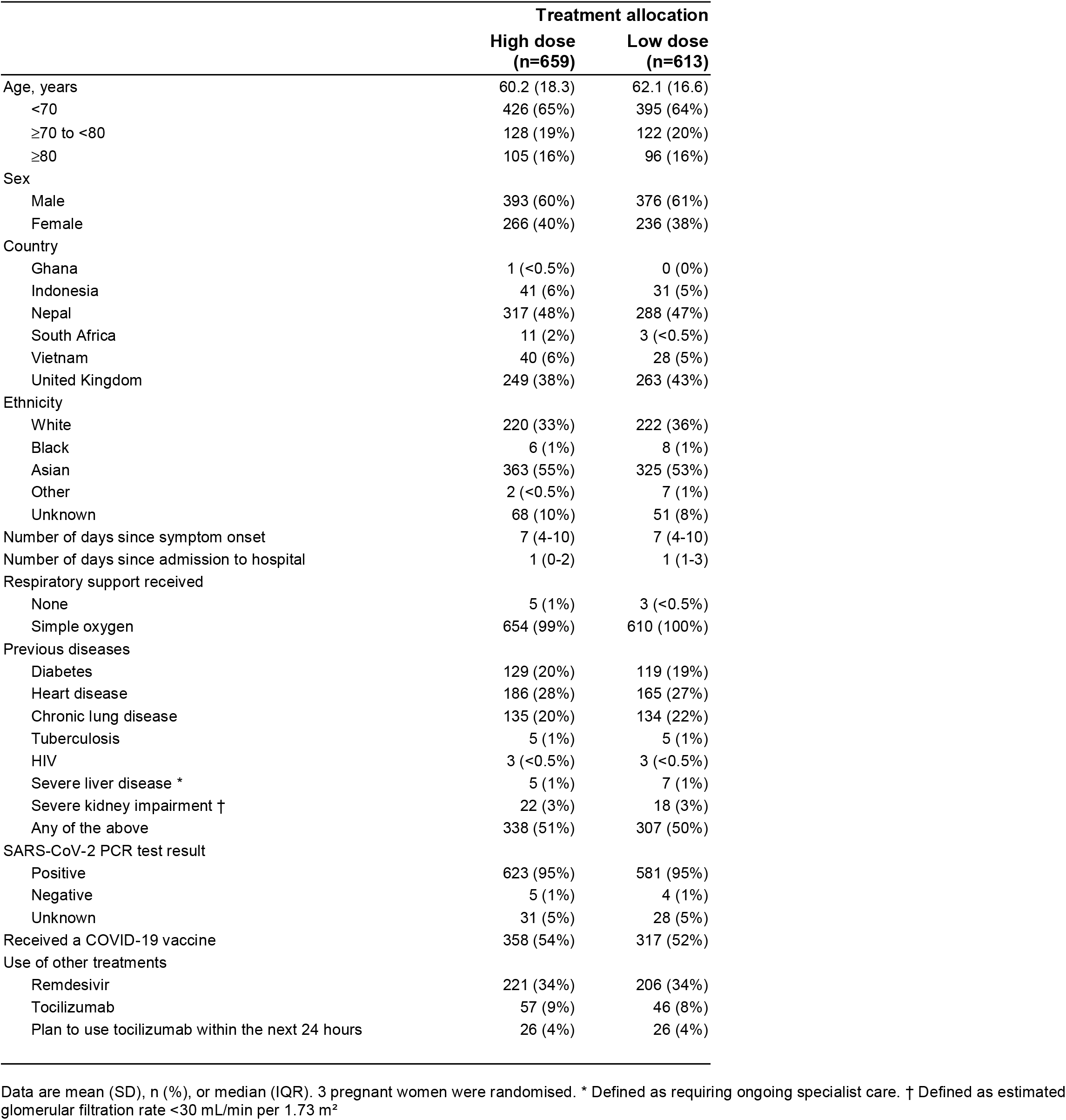
Baseline characteristics by treatment allocation.

|  | Treatment allocation |  |
| --- | --- | --- |
|  | High dose<br>(n=659) | Low dose<br>(n=613) |
| Age, years | 60.2 (18.3) | 62.1 (16.6) |
| <70 | 426 (65%) | 395 (64%) |
| ≥70 to <80 | 128 (19%) | 122 (20%) |
| ≥80 | 105 (16%) | 96 (16%) |
| Sex |  |  |
| Male | 393 (60%) | 376 (61%) |
| Female | 266 (40%) | 236 (38%) |
| Country |  |  |
| Ghana | 1 (<0.5%) | 0 (0%) |
| Indonesia | 41 (6%) | 31 (5%) |
| Nepal | 317 (48%) | 288 (47%) |
| South Africa | 11 (2%) | 3 (<0.5%) |
| Vietnam | 40 (6%) | 28 (5%) |
| United Kingdom | 249 (38%) | 263 (43%) |
| Ethnicity |  |  |
| White | 220 (33%) | 222 (36%) |
| Black | 6 (1%) | 8 (1%) |
| Asian | 363 (55%) | 325 (53%) |
| Other | 2 (<0.5%) | 7 (1%) |
| Unknown | 68 (10%) | 51 (8%) |
| Number of days since symptom onset | 7 (4-10) | 7 (4-10) |
| Number of days since admission to hospital | 1 (0-2) | 1 (1-3) |
| Respiratory support received |  |  |
| None | 5 (1%) | 3 (<0.5%) |
| Simple oxygen | 654 (99%) | 610 (100%) |
| Previous diseases |  |  |
| Diabetes | 129 (20%) | 119 (19%) |
| Heart disease | 186 (28%) | 165 (27%) |
| Chronic lung disease | 135 (20%) | 134 (22%) |
| Tuberculosis | 5 (1%) | 5 (1%) |
| HIV | 3 (<0.5%) | 3 (<0.5%) |
| Severe liver disease * | 5 (1%) | 7 (1%) |
| Severe kidney impairment † | 22 (3%) | 18 (3%) |
| Any of the above | 338 (51%) | 307 (50%) |
| SARS-CoV-2 PCR test result |  |  |
| Positive | 623 (95%) | 581 (95%) |
| Negative | 5 (1%) | 4 (1%) |
| Unknown | 31 (5%) | 28 (5%) |
| Received a COVID-19 vaccine | 358 (54%) | 317 (52%) |
| Use of other treatments |  |  |
| Remdesivir | 221 (34%) | 206 (34%) |
| Tocilizumab | 57 (9%) | 46 (8%) |
| Plan to use tocilizumab within the next 24 hours | 26 (4%) | 26 (4%) |

Data are mean (SD), n (%), or median (IQR). 3 pregnant women were randomised. \* Defined as requiring ongoing specialist care. † Defined as estimated glomerular filtration rate <30 mL/min per 1.73 m<sup>2</sup>

The follow-up form was completed for 657 (99.7%) patients in the higher dose corticosteroid group and 608 (99.2%) patients in the usual care group. Among patients with a completed follow-up form, 91% allocated to higher dose corticosteroid were reported to have received higher dose corticosteroids compared with 1% allocated to usual care (figure 1, webtable 2). Among those with a completed follow-up form allocated usual care, 87% received low dose dexamethasone. Use of other treatments for COVID-19 was broadly similar among patients allocated higher dose corticosteroids and among those allocated usual care, with two-fifths receiving remdesivir and one-tenth receiving an interleukin-6 antagonist during the follow-up period (webtable 2).

Primary and secondary outcome data are known for >99% of randomly assigned patients. Allocation to higher dose corticosteroids was associated with a significant increase in the primary outcome of 28-day mortality compared with usual care alone: 121 (18%) of 659 patients in the higher dose corticosteroid group died vs 75 (12%) of 613 patients in the usual care group (rate ratio 1·56; 95% CI 1·18–2·06; p=0·0020; table 2, figure 2). A similar proportional risk increase was seen in analyses restricted to participants with a positive SARS-CoV-2 PCR test (RR 1.63, 95% CI 1.22-2.17, p=0.0008) or adjusted for baseline age (RR 1.59, 95% CI 1.19-2.12, p=0.0017). In exploratory analyses, the proportional effect of higher dose corticosteroids on mortality was consistent across all 5 pre-specified subgroups (all interaction p-values ≥0.10; Figure 3), including by region of recruitment and by ethnicity.

**Table 2:** Effect of allocation to higher dose corticosteroid on key study outcomes.

|  | Treatment allocation |  | RR (95% CI) |
| --- | --- | --- | --- |
|  | High dose<br>steroids<br>(n=659) | Usual care<br>(n=613) |  |
| **Primary outcome** |  |  |  |
| 28-day mortality | 121 (18.4%) | 75 (12.2%) | 1.56 (1.18-2.06) |
| **Secondary and other key outcomes** |  |  |  |
| Median duration of hospitalization, days | 9 (5 to 17) | 9 (5 to 16) |  |
| Discharged from hospital alive within 28 days | 527 (80.0%) | 505 (82.4%) | 0.92 (0.81-1.04) |
| Invasive mechanical ventilation or death within 28 days | 129 (19.6%) | 80 (13.1%) | 1.50 (1.16-1.94) |
| Use of ventilation | 119 (18.1%) | 84 (13.7%) | 1.32 (1.02-1.70) |
| Non-invasive ventilation | 108 (16.4%) | 82 (13.4%) | 1.23 (0.94-1.60) |
| Invasive mechanical ventilation | 22 (3.3%) | 14 (2.3%) | 1.46 (0.75-2.83) |
| Renal replacement therapy (RRT) * | 12/659 (1.8%) | 8/613 (1.3%) | 1.40 (0.57-3.39) |

Data are n (%) or n/N (%), unless otherwise indicated. RR=rate ratio for the outcomes of 28-day mortality and hospital discharge, and risk ratio for other outcomes. CI=confidence interval. \* Analyses exclude those on haemodialysis or haemofiltration at randomisation.

**Figure 2:**
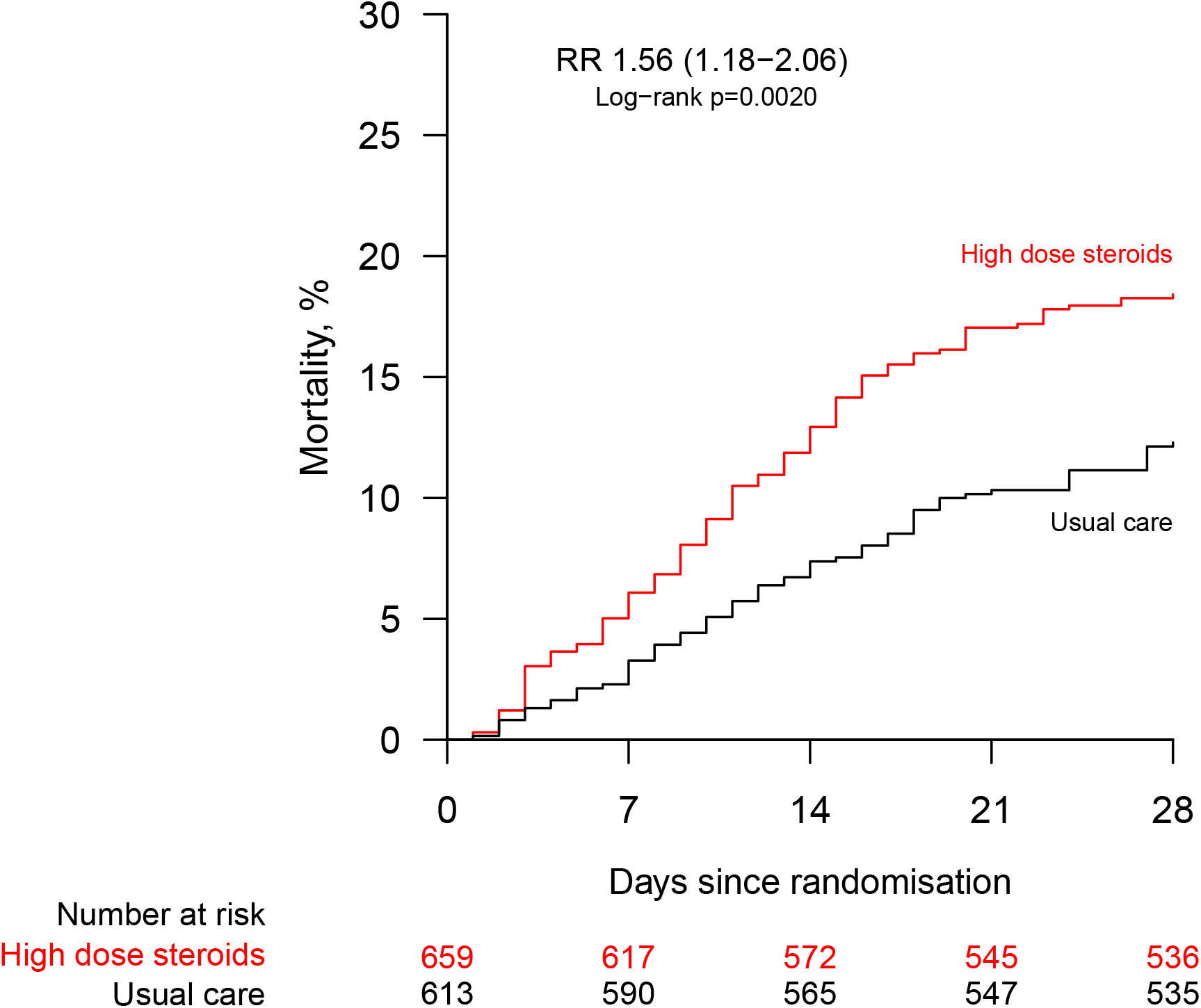
Effect of allocation to higher dose corticosteroids or usual care (lower dose corticosteroids) on 28-day mortality in patients receiving no oxygen or simple oxygen only. RR = rate ratio

**Figure 3:**
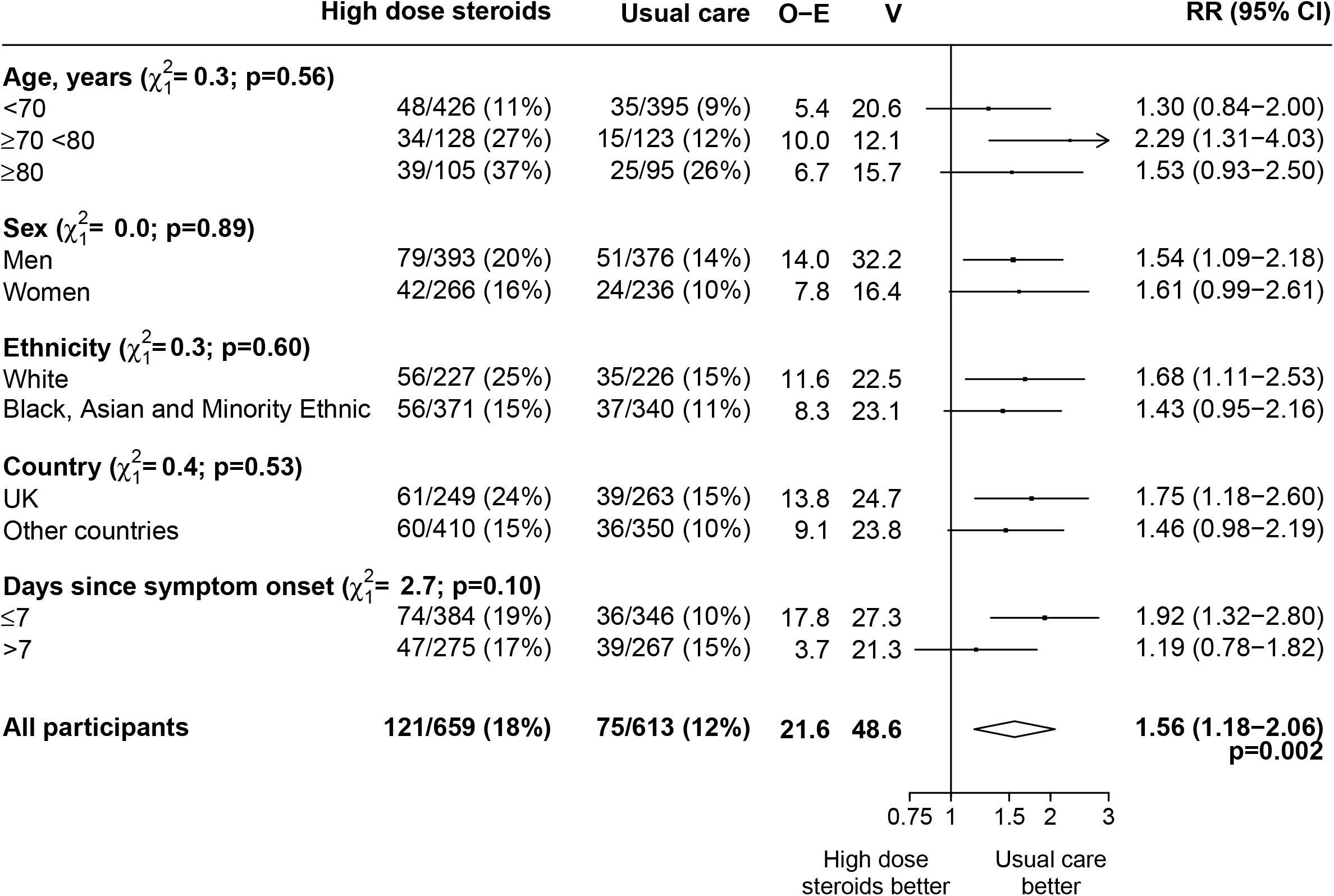
Effect of allocation to higher dose corticosteroids or usual care (lower dose corticosteroids) on 28-day mortality in patients receiving no oxygen or simple oxygen only by other baseline characteristics. Subgroup−specific rate ratio estimates are represented by squares (with areas of the squares proportional to the amount of statistical information) and the lines through them correspond to the 95% CIs. The days since onset subgroup excludes those with missing data, but these patients are included in the overall summary diamond. RR=rate ratio.

Discharge alive within 28 days was similar among those allocated to higher dose corticosteroids compared with usual care (80% vs. 82%; rate ratio 0·92, 95% CI 0·81 to 1.04; median 9 days [IQR 5 to 17] vs. 9 days [IQR 5 to 16]) (table 2; webfigure 1). Allocation to higher dose corticosteroids was associated with a higher risk of progressing to the composite secondary outcome of invasive mechanical ventilation or death (20% vs. 13%, risk ratio 1·50, 95% CI 1·16 to 1·94) (table 2; webfigure 2). There were no significant differences in use of ventilation or receipt of haemodialysis or haemofiltration (table 2).

Three quarters of the deaths within 28 days were attributed to COVID-19 (webtable 3). Allocation to higher dose corticosteroids was associated with an increase in pneumonia reported as not due to COVID-19 (10% vs. 6%, absolute risk increase 3.7%, 95% CI 0.7 to 6.6; webtable 4), but there were no significant differences in the rates of other presentations of non-coronavirus infection, new onset cardiac arrhythmia, thrombotic events, or clinically significant bleeding (webtable 4). Allocation to higher dose corticosteroids was associated with an increase in clinically significant hyperglycaemia (ketoacidosis, hyperglycaemic hyperosmolar state, or hyperglycaemia requiring new use of insulin) occurring in 22% vs. 14% participants (absolute risk increase 7.4%, 95% CI 3.2 to 11.5) (webtable 4). There were 37 reports (among 24 participants) of a serious adverse reaction believed to be related to treatment with higher dose corticosteroids (webtable 5), including 15 reports of a serious non-COVID infection, 9 with hyperglycaemia and 5 with gastro-intestinal bleeding.

## DISCUSSION

In this large, randomised trial, allocation to higher dose corticosteroids significantly increased 28-day mortality for those patients with hypoxia requiring simple oxygen (or no oxygen). There was also an increase in the risk of pneumonia due to non-COVID-19 causes and in hyperglycaemic episodes requiring new use of insulin. This hazard appeared similar among patients from the UK and from Asia and Africa, where the prevalence of TB, HIV and other infections is greater. Within five days of receiving the DMC recommendation regarding the hazard associated with higher dose corticosteroids in this patient group, we had notified all investigators and relevant regulatory authorities and ethics committees, including the US Food and Drugs Administration, the European Medicines Agency, and the World Health Organization, and made the information publicly available via our public website.

Strengths of the RECOVERY trial are that it is randomised, has a large sample size, broad eligibility criteria and more than 99% of patients in this analysis have been followed up for the primary outcome. It was also conducted in areas with high (South and Southeast Asia, Africa) and low (UK) prevalence of HIV, TB, and other infections. The study has some limitations: this randomised trial is open label (i.e., participants and local hospital staff are aware of the assigned treatment), however, the outcome of death is unambiguous. Information on radiological, virological or physiological outcomes was not collected. Only about 10% of participants in this evaluation of high-dose corticosteroids received an interleukin-6 antagonist or baricitinib so we are unable to assess any possible interactions between corticosteroid dose and other immunomodulatory treatments. The enrolment of patients on no oxygen or simple oxygen was closed early on the basis of an interim analysis by the independent Data Monitoring Committee. Consequently, it is possible that the size of the hazards presented may be an over-estimate of the true effects of higher dose corticosteroids.^22^

Prior to the COVID-19 pandemic, the role of corticosteroids in acute lung injury was unclear.^23^ This began to change when the RECOVERY trial showed a clear reduction in 28 days mortality with the use of 6mg dexamethasone once daily for up to 10 days in COVID-19 patients requiring oxygen therapy.^1^ The benefit of corticosteroids amongst critically ill COVID-19 patients was subsequently confirmed in other randomized controlled trials and in a meta-analysis.^9,24^ A broader meta-analysis of the effects of corticosteroids in both COVID-related ARDS and non-COVID ARDS has reported that corticosteroids probably reduce mortality in patients with ARDS regardless of the aetiology.^25^

Previous difficulties in determining the role of corticosteroids in acute lung injury may have arisen because the benefits and hazards of their use may vary by dose, co-morbidities, and the severity of lung injury (which is reflected in the level of respiratory support required).^25,26^ The initial dexamethasone result from RECOVERY showed a clear difference in effect of low-dose corticosteroids by disease severity (p value for trend <0.001), with the greatest benefit seen in the most severely ill. 28-day mortality was reduced by about one-third in those requiring invasive mechanical ventilation or ECMO, with a more modest reduction of one-fifth in those requiring simple oxygen or non-invasive ventilation, and no benefit observed in those not requiring oxygen.^1^

The results we present in this paper show that in hypoxic COVID-19 patients who require only simply oxygen therapy, a higher dose of corticosteroids (dexamethasone 20 mg once daily for 5 days followed by 10 mg once daily for 5 days or until discharge if sooner) is harmful compared to usual care (which included use of lower dose corticosteroids). However, it remains an open question whether increasing the dose of corticosteroids above 6mg dexamethasone per day is helpful for COVID-19 patients requiring non-invasive or invasive mechanical ventilation – the RECOVERY trial continues to seek an answer for that question.

In summary, this large, randomised trial demonstrates that, compared to standard dose corticosteroids, use of higher dose corticosteroids increases mortality for hypoxic patients with COVID-19 who are not receiving non-invasive or invasive mechanical ventilation.

## Research in context

### Evidence before this study

We searched MEDLINE, Embase, MexRxiv and the WHO International Clinical Trials Registry Platform between Sept 1, 2019, and Nov 23, 2022 for randomised controlled trials comparing the effect of different doses of systemic corticosteroids in patients hospitalised with COVID-19 receiving no or simple oxygen at randomisation, using the search terms (Coronavirus infection OR SARS-CoV-2 OR SARS-CoV2 OR SARSCoV2 OR COVID OR COVID-19 OR COVID19 OR 2019-nCoV OR Coronavirus or Coronavirinae) AND (corticosteroid OR dexamethasone OR glucocorticoid OR steroid OR hydrocortisone OR methylprednisolone OR prednisolone OR betamethasone) and using validated filters to select for randomised controlled trials. No language restrictions were applied.

We identified twelve relevant randomised trials with results available that assessed different doses of corticosteroids in hospitalised COVID-19 patients, at least some of whom were receiving simple oxygen therapy at randomisation: six assessed higher dose dexamethasone (12-24 mg per day) and six assessed methylprednisolone (60-1000 mg per day), all compared to lower dose dexamethasone (6-8 mg per day).^27-38^ Four of the trials included only a non-critically ill population and the remainder included a mixed illness severity population.^28,31,37,38^ Eleven of the trials have been fully published of which five were considered to have low risk of bias for the 28-day mortality outcome and six having some concerns (including lack of information about randomisation process, lack of information about prespecified analyses, crossover between randomised groups and post-randomisation exclusion of patients in the analysis population) (appendix p31). All but one of the trials have found no statistically significant difference in mortality between the treatment groups. The exception was a small trial comparing methylprednisolone (2mg/kg/day for 3 days then 1mg/kg/day for 3 days) with dexamethasone (8mg daily for 10 days) that included 140 patients and 18 deaths, that reported that methylprednisolone was associated with significantly improved mortality at 10 days.^38^ There was no information in the trial report about the randomisation process or whether the trial was analysed as pre-specified, conferring some concerns over risk of bias.

### Added value of this study

The Randomised Evaluation of COVID-19 Therapy (RECOVERY) trial is the largest randomised trial of the effect of different doses of corticosteroids in patients hospitalised with COVID-19 and included patients from 3 continents. We found that among hypoxic patients on simple or no oxygen, randomisation to higher dose corticosteroids (dexamethasone 20 mg daily for 5 days followed by dexamethasone 10 mg for 5 days, or until discharge if sooner) vs. usual care (which included dexamethasone 6 mg once daily in 87% of participants) resulted in an increased risk of all-cause mortality, Overall, 121 (18%) of 659 patients allocated to higher dose corticosteroids versus 75 (12%) of 613 patients allocated to usual care died within 28 days (rate ratio [RR] 1·56; 95% CI 1·18-2·06; p=0·0020). There was also an excess of pneumonia reported to be due to non-COVID infection (10% vs. 6%; absolute difference 3.7%; 95% CI 0.7-6.6) and an increase in hyperglycaemia requiring increased insulin dose (22% vs. 14%; absolute difference 7.4%; 95% CI 3.2-11.5).

### Implications of all the available evidence

Among hospitalised patients with COVID-19 who require oxygen or ventilatory support, low-dose corticosteroids reduce the risk of death. However, among those requiring simple oxygen only, higher doses of corticosteroids increase the risk of death compared to low-dose corticosteroids. It remains unclear whether using a higher dose of corticosteroids is beneficial among those requiring non-invasive or invasive ventilation – the RECOVERY trial continues to study this question.

## Supporting information

CONSORT check-list

## Data Availability

The protocol, consent form, statistical analysis plan, definition & derivation of clinical characteristics & outcomes, training materials, regulatory documents, and other relevant study materials are available online at www.recoverytrial.net. As described in the protocol, the Trial Steering Committee will facilitate the use of the study data and approval will not be unreasonably withheld. Deidentified participant data will be made available to bona fide researchers registered with an appropriate institution within 3 months of publication. However, the Steering Committee will need to be satisfied that any proposed publication is of high quality, honours the commitments made to the study participants in the consent documentation and ethical approvals, and is compliant with relevant legal and regulatory requirements (e.g. relating to data protection and privacy). The Steering Committee will have the right to review and comment on any draft manuscripts prior to publication. Data will be made available in line with the policy and procedures described at: https://www.ndph.ox.ac.uk/data-access. Those wishing to request access should complete the form at
https://www.ndph.ox.ac.uk/files/about/data_access_enquiry_form_13_6_2019.docx
and e-mailed to

## Contributors

This manuscript was initially drafted by the PWH and MJL, further developed by the Writing Committee, and approved by all members of the Trial Steering Committee. PWH and MJL vouch for the data and analyses, and for the fidelity of this report to the study protocol and data analysis plan. PWH, BB, RLH, JA, JKB, MB, LCC, JD, SNF, TJ, EJ, KJ, WSL, MM, AMo, AMu, KR, GT, RH, and MJL designed the trial and study protocol. MM, LP, MC, G P-A, EK, MR, RKJ, NTP, US, DP, PNT, NN, ES, LM, RS, and the Data Linkage team at the RECOVERY Coordinating Centre, and the Health Records and Local Clinical Centre staff listed in the appendix collected the data. NS and JRE had access to the study data and did the statistical analysis. All authors contributed to data interpretation and critical review and revision of the manuscript. PWH and MJL had access to the study data and had final responsibility for the decision to submit for publication.

## Writing Committee (on behalf of the RECOVERY Collaborative Group)

Peter W Horby,* Jonathan R Emberson,* Buddha Basnyat,* Mark Campbell, Leon Peto, Guilherme Pessoa-Amorim, Natalie Staplin, Raph L Hamers, John Amuasi, Jeremy Nel, Evelyne Kestelyn, Manisha Rawal, Roshan Kumar Jha, Nguyen Thanh Phong, Uun Samardi, Damodar Paudel, Pham Ngoc Thach, Nasronudin Nasronudin, Emma Stratton, Louise Mew, Rahul Sarkar, J Kenneth Baillie, Maya H Buch, Jeremy Day, Saul N Faust, Thomas Jaki, Katie Jeffery, Edmund Juszczak, Marian Knight, Wei Shen Lim, Marion Mafham, Alan Montgomery, Andrew Mumford, Kathryn Rowan, Guy Thwaites,^+^ Richard Haynes,^+^ Martin J Landray.^+ *^

PWH, JRE and BB made an equal contribution ^+^

GT, RH and MJL made an equal contribution

## Data Monitoring Committee

Peter Sandercock, Janet Darbyshire, David DeMets, Robert Fowler, David Lalloo, Mohammed Munavvar, Adilia Warris, Janet Wittes.

## Declaration of interests

The authors have no conflict of interest or financial relationships relevant to the submitted work to disclose. No form of payment was given to anyone to produce the manuscript. All authors have completed and submitted the ICMJE Form for Disclosure of Potential Conflicts of Interest. The Nuffield Department of Population Health at the University of Oxford has a staff policy of not accepting honoraria or consultancy fees directly or indirectly from industry (see https://www.ndph.ox.ac.uk/files/about/ndph-independence-of-research-policy-jun-20.pdf).

## Data sharing

The protocol, consent form, statistical analysis plan, definition & derivation of clinical characteristics & outcomes, training materials, regulatory documents, and other relevant study materials are available online at www.recoverytrial.net. As described in the protocol, the Trial Steering Committee will facilitate the use of the study data and approval will not be unreasonably withheld. Deidentified participant data will be made available to bona fide researchers registered with an appropriate institution within 3 months of publication. However, the Steering Committee will need to be satisfied that any proposed publication is of high quality, honours the commitments made to the study participants in the consent documentation and ethical approvals, and is compliant with relevant legal and regulatory requirements (e.g. relating to data protection and privacy). The Steering Committee will have the right to review and comment on any draft manuscripts prior to publication. Data will be made available in line with the policy and procedures described at: https://www.ndph.ox.ac.uk/data-access. Those wishing to request access should complete the form at https://www.ndph.ox.ac.uk/files/about/data_access_enquiry_form_13_6_2019.docx and e-mailed to

## Acknowledgements

Above all, we would like to thank the thousands of patients who participated in this trial. We would also like to thank the many doctors, nurses, pharmacists, other allied health professionals, and research administrators at participating hospital organisations. Supported in the UK by staff at the National Institute of Health and Care Research (NIHR) Clinical Research Network, NHS DigiTrials, Public Health England, Department of Health & Social Care, the Intensive Care National Audit & Research Centre, Public Health Scotland, National Records Service of Scotland, the Secure Anonymised Information Linkage (SAIL) at University of Swansea, and the NHS in England, Scotland, Wales and Northern Ireland.

The RECOVERY trial is supported by grants to the University of Oxford from□UK Research and Innovation (UKRI) and NIHR (MC_PC_19056), the Wellcome Trust (Grant Ref: 222406/Z/20/Z) through the COVID-19 Therapeutics Accelerator, and by core funding provided by□the NIHR Oxford Biomedical Research Centre, the Wellcome Trust,□the□Bill and Melinda Gates Foundation, the□Foreign, Commonwealth and Development Office, Health Data Research UK, the□Medical Research Council Population Health Research Unit, the NIHR Health Protection Unit in Emerging and Zoonotic Infections, and□NIHR Clinical Trials Unit Support Funding. TJ is supported by a grant from UK Medical Research Council (MC_UU_00002/14). WSL is supported by core funding provided by NIHR Nottingham Biomedical Research Centre. Tocilizumab, casirivimab and imdevimab, sotrovimab, and empagliflozin were provided through support from Roche, Regeneron, GSK, and Boehringer Ingelheim, respectively. Colchicine for use in Indonesia was provided by Combiphar. The views expressed in this publication are those of the authors and not necessarily those of the NHS, the NIHR or the Department of Health and Social Care. For the purpose of Open Access, the author has applied a CC BY public copyright licence to any Author Accepted Manuscript version arising from this submission.

## Notes

### Competing Interest Statement

The authors have declared no competing interest.

### Clinical Trial

The RECOVERY trial is registered with ISRCTN (50189673) and clinicaltrials.gov (NCT04381936).

### Clinical Protocols

https://www.recoverytrial.net

### Author Declarations

The trial is conducted in accordance with the Principles of the International Conference on Harmonisation-Good Clinical Practice guidelines and approved by the UK Medicines and Healthcare products Regulatory Agency (MHRA) and the Cambridge East Research Ethics Committee (ref: 20/EE/0101). Further details are listed in the supplementary appendix.

